# Silent Cries Across Borders: Determinants of Suicidal Ideation and Attempts in Sub-Saharan Africa - Evidence from WHO STEPS Surveys (2014–2019)

**DOI:** 10.1101/2025.09.09.25335473

**Authors:** Ilunga Ngalande, Adoration Chigere, Wingston Ng’ambi, Cosmas Zyambo

## Abstract

**Introduction:** Suicide is a major global health concern, claiming approximately 800,000 lives annually, predominantly in low- and middle-income countries. In Sub-Saharan Africa, research on suicidal behavior remains limited, hindered by stigma, inadequate mental health infrastructure and data scarcity. This study examines the prevalence and predictors of suicidal ideation and attempts across 12 African nations to guide culturally sensitive prevention strategies.

**Methods:** This cross-sectional study analyzed secondary data from WHO STEPS surveys (2014-2019), the study employed a multistage cluster sampling and standardized assessment to evaluate suicidal behavior among individuals aged 15 and older. A total of 14,374 participants were assessed for ideation and 16,041 for attempts. Multivariable logistic regression and spatial mapping were used to identify risk factors and geographic trends.

**Results:** Finding revealed a 6.34% prevalence of suicidal ideation and a 5.69% for suicidal attempts, with higher rates among females and youths aged 15–34. Males were significantly less likely to report suicidal thoughts than females (AOR = 0.61;95% CI: 0.52–0.72; *p* < 0.001). After adjustment, men were approximately 39% less likely to attempt suicide than women (AOR = 0.61, 95% CI: 0.52–0.72; *p* < 0.001). Key risk factors included gender, country of residence (Eswatini, Malawi, or Zambia), hypertension, high salt intake, and alcohol use. Alcohol consumption was notably linked to a two-ford increase in both ideation and attempts. Variables such as education, marital status, and urban-residence showed minimal influence after adjustment.

**Conclusion:** This study underscores the urgent need to integrate mental health services into primary care systems, enhance public awareness, and reduce stigma surrounding suicide. Expanding access to affordable, culturally appropriate mental health resources is critical to addressing the high burden of suicidal behavior in Sub-Saharan Africa.

## INTRODUCTION

Suicide is a major global health crisis, responsible for an estimated 800,000 deaths annually, with 79% occurring in low- and middle-income countries (LMICs) (1). According to the World Health Organization (WHO), for every suicide, there are approximately 20 attempts, reflecting the broader burden of suicidal behaviors worldwide (2). Although global suicide rates declined by around 36% between 2000 and 2019, this trend has not been uniform, with certain vulnerable populations continuing to face disproportionately high rates of suicidal ideation and attempts (3). Notably, several Sub-Saharan African (SSA) countries have exhibited concerning increases, with suicidal ideation affecting an estimated 10–15% of adults (4,5). While established risk factors such as mental health disorders, substance use, chronic illness, and socioeconomic adversity are widely recognized, their expression and impact often vary across cultural and geographic contexts (6,7).

In SSA, the true burden of suicidal behaviors remains inadequately understood. Reported suicide rates in the region range from 3.2 to 34.6 per 100,000 population, reflecting both genuine epidemiological variation and significant challenges in data collection, stigma, and underdeveloped surveillance systems (8,9). SSA faces unique contextual pressures including rapid urbanization, high unemployment, and severely limited mental health resources, with only 0.9 mental health workers per 100,000 population compared to a global average of 9 (10,11). Cultural taboos surrounding mental illness and suicide further hinder recognition and intervention. Additionally, region-specific stressors such as high HIV/AIDS prevalence, political instability, and food insecurity may interact with global suicide risk factors but remain under examined in existing literature (12). These challenges, compounded by systemic data gaps and chronic underachievement in mental health infrastructure, contribute to the region’s under representation in global suicide research and limit effective policy response (13,14).

The WHO STEPwise Approach to Surveillance (STEPS) provides a valuable yet underutilized resource for addressing critical knowledge gaps on suicidal behaviors in SSA. Conducted between 2014 and 2019 in several SSA countries, these nationally representative surveys offer harmonized data on a broad range of non-communicable disease risk factors, including mental health indicators such as suicidal ideation and suicide attempts (15). The standardized STEPS methodology enables valid cross-country comparisons while capturing detailed contextual data on socio-demographic characteristics, behavioral risk factors, chronic conditions, and health outcomes (16). This makes STEPS uniquely suited for population-level analyses of suicide-related outcomes across diverse SSA settings, facilitating a deeper understanding of both common and context-specific drivers of suicidal behavior (17).

Despite the potential of the STEPS data set, no multi-country analysis has yet explored the prevalence and risk factors of suicidal behaviors in Sub-Saharan Africa using this resource. This study addresses that gap by analyzing the most recent WHO STEPS survey data (2014–2019) from multiple SSA countries. The aim of the paper is to estimate the prevalence and the determinants of suicidal ideation and suicide attempts among adults in the Sub-Saharan Africa. The findings will inform the development of culturally relevant, evidence-based suicide prevention strategies in SSA. By contributing critical data to a historically under-researched area, this study advances global health efforts to reduce premature mortality from mental health conditions, in line with Sustainable Development Goal 3 and 4 (18).

## METHODS

### Study Design and Setting

This study used a cross-sectional analytical design based on secondary data from WHO STEPwise Approach to Surveillance (STEPS) surveys conducted between 2014 and 2019 across multiple Sub-Saharan African (SSA) countries.(19). The STEPS surveys are nationally representative, population-based assessments designed to collect standardized data on non-communicable disease (NCD) risk factors, including mental health indicators such as suicidal ideation and suicide attempts (20). Utilizing a multistage cluster sampling approach, the surveys target adults aged 18–69 years, ensuring broad demographic and geographic representation within each participating country (21).

Data collection took place across diverse Sub-Saharan African (SSA) settings, encompassing both urban and rural areas with varied socioeconomic and cultural contexts. Countries were included in the analysis based on the availability of recent STEPS survey data that incorporated modules on suicidal behaviors. The standardized STEPS methodology ensures comparability of findings across countries, despite differences in health infrastructure and reporting systems (22,23). This multi-country dateset offers a robust foundation for examining the prevalence, risk factors, and protective factors associated with suicidal behaviors among adult populations in SSA (24).

### Data management

This study utilized data from WHO STEPS surveys conducted between 2014 and 2019 across 12 Sub-Saharan and North African countries: Algeria, Benin, Botswana, Eswatini, Ethiopia, Kenya, Malawi, Morocco, São Tomé and Príncipe (STP), Sudan, Uganda, and Zambia. These nationally representative datasets were obtained from the WHO NCD Micro-data Repository and harmonized for multi-country analysis (25).

The data management process involved standardizing variable names and coding schemes to ensure consistency across countries. Key socio-demographic and behavioral variables were extracted, including age (categorized as 18–29, 30–44, and 45–69 years), sex (male, female), residence (urban, rural), education level (no formal education, primary, secondary, tertiary), and marital status (currently married, never married, formerly married). Country identifiers were retained for both stratified and pooled analyses. Behavioral and clinical risk factors included current smoking (yes/no), harmful alcohol use (defined according to WHO guidelines), low fruit and vegetable intake (fewer than five servings per day), hypertension (HTN) (based on blood pressure readings or self-reported diagnosis), and diabetes mellitus (DBM) (based on fasting blood glucose or self-report).

The primary outcomes of interest in this study were suicidal ideation and suicide attempts, as self-reported in the WHO STEPS surveys. Suicidal ideation was defined by an affirmative response to the question: *’Have you ever seriously considered attempting suicide during the past 12 months?’* Suicide attempts were identified through the question: *’During the past 12 months, have you attempted suicide?’* Both outcomes were treated as binary variables (Yes/No). Participants who responded affirmatively were classified as having experienced suicidal ideation or attempted suicide, respectively. These outcomes were analyzed independently and, in exploratory analyses, combined into a composite indicator of suicidal behavior to examine shared and distinct risk factor profiles. While self-report data may be influenced by stigma and under reporting, the confidential administration of STEPS surveys and supporting literature validate their utility in population-based mental health research. These standardized outcome measures enabled robust cross-country comparisons and multi-variable modeling of associations with socio-demographic, behavioral, and clinical determinants across the 12 participating African countries.

Data cleaning and management were performed using R (version 4.3.2). Missing values for key variables were assessed and addressed through listwise deletion where appropriate. Sensitivity analyses were conducted to evaluate the robustness of findings to different handling strategies for missing data. Each country-specific dataset was individually reviewed for quality and completeness prior to merging. The final pooled dataset was used to conduct descriptive analyses, multivariable logistic regression, and spatial analyses to examine associations between identified determinants and suicidal ideation or suicide attempts across SSA.

### Data Analysis

All statistical analyses were conducted using R (version 4.3.2). Data were cleaned, harmonized, and merged across 12 participating countries: Algeria, Benin, Botswana, Eswatini, Ethiopia, Kenya, Malawi, Morocco, São Tomé and Príncipe, Sudan, Uganda, and Zambia. Sampling weights provided by the WHO STEPS surveys were applied to all analyses to account for the complex survey design and ensure national representativeness (26).

Descriptive statistics were used to summarize socio-demographic and behavioral characteristics of the study population (27). Continuous variables were reported as means with standard deviations or medians with inter-quartile ranges, depending on distribution. Categorical variables were summarized using weighted proportions and 95% confidence intervals. Country-level prevalence estimates of suicidal ideation and suicide attempts were calculated and disaggregated by sex, age group, and urban–rural residence to explore population-level patterns.

Bivariate analyses were conducted using Rao-Scott chi-squared tests and survey-weighted logistic regression models to assess associations between each potential determinant and the primary outcomes—suicidal ideation and suicide attempts. Independent variables included age (categorized), sex, place of residence (urban/rural), marital status, educational attainment, current smoking, harmful alcohol use, hypertension, diabetes, and inadequate fruit and vegetable intake.

Multivariable logistic regression models were subsequently fitted to identify factors independently associated with each outcome, adjusting for all covariates identified a priori and those significant at *p* < 0.2 in bivariate analyses. Adjusted odds ratios (AORs) with 95% confidence intervals were reported. Multi-collinearity among predictors was assessed using standard collinearity diagnostics.

Spatial analysis was performed to explore geographic variation in the prevalence of suicidal behaviors, incorporating choropleth mapping and district-level correlations with indicators such as chronic disease burden (e.g., hypertension, diabetes) and household-level illness (number of reported sick adults and children) (28). All statistical tests were two-sided, with significance set at *p* < 0.05.

### Ethical considerations

Ethical approval for this study was obtained from the University of Zambia Biomedical Research Ethics Committee (UNZABREC; Ref. No. 6114-2024) and the National Health Research Authority (NHRA) of Zambia (Ref. No. NHRAR-R-2142/31/10/2024). Permission to use the secondary data was granted by the World Health Organization (WHO), which oversees the administration of the STEPS surveys. All participants provided informed consent prior participation in written form. The datasets were accessed via the WHO NCD Micro-data Repository. In accordance with ethical standards for research involving human participants, all data were fully anonymized prior to analysis. No personal identifiers were included, and only coded responses were used to ensure participant privacy. Data were handled in strict compliance with international ethical guidelines and national regulatory requirements to maintain confidentiality and data security.

## RESULTS

### Characteristics and prevalence of respondents on suicidal ideation in Algerian, Eswatini, Malawi and Zambia from 2014-2019

The analysis included a weighted sample of 14,374 individuals aged 15 years and older, with an approximately equal gender distribution (50.56% female, 49.44% male). The majority of participants were between 20 and 44 years of age, with the highest representation in the 20–24 (16.36%) and 25–29 (15.94%) age groups. Just over half of the sample resided in urban areas (52.11%). In terms of educational attainment, most respondents had completed either primary (45.54%) or secondary (32.01%) education. Regarding employment status, the largest proportion were either unpaid workers or retirees (45.83%), followed by self-employed individuals (30.09%). Marital status data indicated that 65.48% of participants were currently married, while approximately 30% had never been married. Geographically, the sample was predominantly drawn from Algeria (62.56%), followed by Malawi (19.13%) and Zambia (17.55%), with Eswatini contributing a smaller proportion (0.76%). Most participants did not report a diagnosis of diabetes (96.49%) or hypertension (88.45%).In terms of health behaviors, a substantial proportion of participants reported poor dietary habits: 71.43% consumed inadequate amounts of fruit, and 28.27% had low vegetable intake. Additionally, 78.61% did not meet the recommended levels of physical activity. Smoking and alcohol use varied across the sample, with 14.58% identifying as current smokers and 2.61% reporting alcohol consumption. Data collection occurred across three survey years: 2014, 2016, and 2017. The overall prevalence of suicidal ideation among the study population was 6.34% (n = 911), indicating that a significant minority of individuals reported experiencing suicidal thoughts a critical public mental health concern in the sampled countries.

Females reported a higher prevalence of suicidal ideation (6.06%) compared to males (3.47%). Age-specific prevalence was highest among adolescents aged 15–19 years (5.16%) and adults aged 20–34 years (ranging from 4.47% to 5.85%), indicating a concentration of suicidal thoughts among younger populations. An inverse relationship was observed with educational attainment: individuals with no formal education (4.77%) or only primary education (5.59%) reported higher prevalence rates, while those with tertiary education had the lowest (2.53%). Among occupational groups, unpaid or retired individuals exhibited the highest prevalence (5.51%). Similarly, formerly married individuals had the highest prevalence across marital status categories (9.42%).

Geographic and biomedical factors also showed variation. Rural residents reported a slightly higher prevalence (5.28%) than urban residents (4.32%). Individuals with diabetes had a higher prevalence of suicidal ideation (6.80%) compared to those without diabetes (4.52%). Suicidal ideation was also more common among participants with high salt intake (5.63%), low fruit (5.01%) and low vegetable consumption (5.09%). Behavioral risk factors were significantly associated with suicidal ideation. Current smokers had a higher prevalence (5.93%) than never-smokers (4.84%), while individuals who reported alcohol use had nearly double the prevalence (10.88%) compared to non-drinkers (4.62%). Prevalence of suicidal ideation also varied by survey year: it peaked in 2014 (9.16%), declined in 2016 (3.16%), and rose again in 2017 (7.46%). These findings highlight the multifaceted nature of suicidal ideation and its associations with socio-demographic, behavioral, and health-related factors.

### Factors associated with suicidal ideation in Algerian, Eswatini, Malawi and Zambia on suicidal attempts from 2014-2019

Gender emerged as a significant predictor of suicidal ideation. Males were significantly less likely to report suicidal thoughts than females, with an adjusted odds ratio (AOR) of 0.61 (95% CI: 0.52–0.72; *p* < 0.001), suggesting that women may experience unique psychological, social, or economic stressors that elevate their risk. Age, however, did not show a consistent pattern. Compared to adolescents aged 15–19 years (reference group), none of the older age groups demonstrated statistically significant adjusted odds of suicidal ideation, indicating that age may not independently predict risk once other covariates are accounted for. While marital status and occupational category appeared significant in crude analyses, these associations attenuated after adjustment. Formerly married individuals initially exhibited more than twice the odds of suicidal ideation (Crude OR = 2.04), but this association was no longer significant in the multivariable model (AOR = 1.18; *p* = 0.23). Similarly, higher crude odds observed among non-paid/retired and self-employed individuals did not persist after controlling for confounders. These findings suggest that the apparent associations with marital and occupational status may be mediated by other socio-demographic or health-related factors. Country of residence was a strong predictor of suicidal ideation. Compared to individuals in Algeria (reference category), participants from Eswatini, Malawi, and Zambia had significantly higher adjusted odds of reporting suicidal thoughts. Eswatini exhibited the highest risk (AOR = 3.24, 95% CI: 2.61–4.02), followed by Zambia (AOR = 2.66) and Malawi (AOR = 2.29), all with *p* < 0.001. These differences likely reflect underlying disparities in national mental health infrastructure, socioeconomic stressors, and broader political or health system vulnerabilities.

Urban versus rural residence was not significantly associated with suicidal ideation after adjustment, although crude analyses showed slightly elevated odds among rural residents. This suggests that geographic location alone does not independently predict risk once individual-level factors are considered. Educational attainment demonstrated some protective trends. Individuals with tertiary education had lower crude odds of suicidal ideation (OR = 0.52); however, this association was only marginally significant after adjustment (AOR = 0.72, *p* = 0.07), indicating a potential but inconclusive buffering effect. Primary and secondary education levels were not significantly associated with suicidal ideation when compared to those with no formal education. From a biomedical and behavioral health perspective, hypertension, high salt intake, and alcohol use emerged as significant predictors of suicidal ideation. Individuals with hypertension had 1.5 times higher adjusted odds of reporting suicidal thoughts (AOR = 1.50, 95% CI: 1.25–1.80; *p* < 0.001), suggesting a potential psychosomatic burden or the psychological impact of managing a chronic condition. High salt intake was also significantly associated with suicidal ideation (AOR = 1.38; *p* < 0.001), which may reflect broader patterns of unhealthy dietary behavior and associated stress or comorbidities.

Alcohol use was one of the most prominent behavioral risk factors. Participants who reported alcohol consumption had markedly higher odds of suicidal ideation, with a crude odds ratio of 2.52 (*p* < 0.001), highlighting the strong link between alcohol use and adverse mental health outcomes. In contrast, other behavioral and clinical variables such as fruit and vegetable intake and diabetes status did not show significant associations in adjusted models. These findings indicate that while chronic health conditions and lifestyle factors are important, their influence on suicidal ideation varies, with hypertension, salt intake, and alcohol use standing out as the most impactful biomedical determinants in this analysis.

### Characteristics and prevalence of respondents in Algerian, Eswatini, Malawi and Zambia on suicidal attempts from 2014-2019

The study included 16,041 participants, with an approximately equal gender distribution: 50.56% female and 49.44% male. Participants were predominantly aged 20–34 years, with 16.36% aged 20–24, 15.94% aged 25– 29, and 13.51% aged 30–34. Slightly more than half resided in urban areas (52.11%), while 47.89% lived in rural settings. In terms of education, the majority had completed either primary (45.54%) or secondary (32.01%) education; smaller proportions had no formal education (10.07%) or tertiary education (12.38%). Nearly half were non-paid workers or retired (45.83%), followed by self-employed individuals (30.09%), while government and NGO employees comprised smaller groups. Most participants were currently married (65.48%), with 29.96% never married and 4.56% formerly married.

Geographically, participants were drawn from Algeria (62.56%), Malawi (19.13%), Zambia (17.55%), and Eswatini (0.76%). Biomedical characteristics showed that 3.51% reported having diabetes and 11.55% had hypertension. Regarding health behaviors, 23.71% reported high salt intake, 71.43% had low fruit consumption, and 28.27% reported low vegetable intake, 78.61% did not meet the recommended levels of physical activity. Additionally, 14.58% were current smokers and 2.61% reported alcohol use.

The overall prevalence of suicide attempts was 5.69% (n = 911), while 6.34% of participants reported experiencing suicidal ideation. There was a strong correlation between ideation and attempts, with suicidal ideation emerging as a key predictor of suicide attempts. Alcohol use was also notably associated with increased risk: individuals who consumed alcohol had nearly double the prevalence of suicide attempts (10.88%) compared to non-drinkers (4.62%).

Gender differences were apparent, with females reporting a higher prevalence of suicide attempts (6.06%) than males (3.47%). Suicide attempts were more frequent among younger age groups, particularly those aged 20–24 (5.85%), 15–19 (5.16%), and 30–34 (5.39%). Prevalence was lowest among older adults, especially those aged 60–64 (2.97%) and 55–59 (3.26%).

Rural residents reported a slightly higher prevalence of suicide attempts (5.28%) compared to urban residents (4.32%). Among marital status categories, formerly married individuals had the highest prevalence (9.42%), compared to 4.43% among currently married and 4.84% among never-married participants. Country-level differences were evident, with Malawi and Zambia exhibiting higher rates of suicide attempts relative to Algeria, although exact prevalence figures were not specified.

From a biomedical perspective, individuals with diabetes reported a significantly higher prevalence of suicide attempts (6.8%) compared to those without (4.52%). Similarly, those with hypertension and high salt intake (5.63%) showed elevated risk. Behavioral risk factors also played a role: current smokers reported higher prevalence (5.93%) compared to former smokers (2.64%) and never smokers (4.84%). Low fruit and vegetable intake was modestly associated with higher rates of suicide attempts compared to adequate intake

### Factors associated with suicidal attempts in Algerian, Eswatini, Malawi and Zambia on suicidal attempts from 2014-2019

The analysis indicates that suicide attempts are more strongly associated with contextual (country-level), biological (chronic health conditions), and behavioral (diet and alcohol use) factors than with demographic or socioeconomic characteristics alone. Gender was a significant predictor, males had substantially lower odds of reporting suicide attempts compared to females, a finding consistent across both unadjusted and adjusted models. After adjustment, men were approximately 39% less likely to attempt suicide than women (AOR = 0.61, 95% CI: 0.52–0.72; *p* < 0.001). Age did not show statistically significant associations in adjusted models, although some variability was observed in unadjusted analyses.

Using individuals aged 65 and older as the reference group, no age group demonstrated significantly different adjusted odds, suggesting that age alone is not a strong independent predictor of suicide attempts when controlling for other variables. Marital status and education level also lacked significant associations in adjusted models. While formerly married individuals had higher odds of suicide attempts in crude analysis (OR = 2.04, 95% CI: 1.44–2.91), this association was not significant after adjustment. Similarly, tertiary education initially appeared protective (crude OR = 0.52, *p* = 0.01), but this effect was attenuated in the multivariable model (AOR = 0.72, *p* = 0.07). These results suggest that marital disruption and lower educational attainment may be confounded by other risk factors. Country of residence emerged as a strong contextual determinant. Compared to Algeria, individuals from Eswatini, Malawi, and Zambia had significantly higher odds of reporting suicide attempts, even after adjusting for covariates. Eswatini had the highest adjusted odds (AOR = 3.24, 95% CI: 2.61–4.02; *p* < 0.001), followed by Malawi and Zambia. These findings highlight the influence of broader structural and systemic factors such as economic instability, sociopolitical conditions, and variations in mental health service availability that may drive country-level differences in suicide risk.

Finally, urban versus rural residence was not significantly associated with suicide attempts after adjustment, suggesting that geographic location in itself does not independently influence risk when other individual and contextual factors are accounted for. Biomedical and lifestyle factors were significantly associated with suicide attempts. Individuals with hypertension had 50% higher adjusted odds of attempting suicide (AOR = 1.50, 95% CI: 1.25–1.80; *p* < 0.001), underscoring the potential psychological burden associated with managing chronic illness. High salt intake was also a significant risk factor (AOR = 1.38; *p* < 0.001), which may indicate broader issues related to unhealthy dietary patterns or metabolic stressors linked to poor mental health outcomes. Alcohol use emerged as a particularly concerning behavioral correlate. In crude analysis, individuals who consumed alcohol had more than double the odds of suicide attempts (OR = 2.52; *p* < 0.001), reinforcing its strong association with suicidal behavior.

However, alcohol use was not included in the adjusted model, limiting interpretation of its independent effect and highlighting the need for further research on its causal relationship with suicide risk. Other factors, including diabetes, low fruit and vegetable intake, and smoking history, did not show statistically significant associations in the adjusted models. These findings suggest that while chronic conditions and poor lifestyle behaviors are relevant, not all exert equal influence on suicidal behavior. Hypertension and high salt intake appear to be the most salient biomedical contributors in this analysis.

**TABLE 1:** Characteristics and Prevalence of Respondents that participated in the WHO STEPwise survey for Suicidal Ideation: 2014-2019.

| Characteristics | Total | Suicidal Ideation |  |  |
| --- | --- | --- | --- | --- |
|  | N (%) | No | Yes |  |
| Social-demographic | (14,374) | N | N | % |
| **Age Group** |  |  |  |  |
| 15-19 | 979 (7.31) | 918 | 61 | 5.16 |
| 20-24 | 1943 (16.36) | 1801 | 142 | 5.85 |
| 25-29 | 1891 (15.94) | 1758 | 133 | 4.47 |
| 30-34 | 2007 (13.51) | 1861 | 146 | 5.39 |
| 35-39 | 1899 (12.63) | 1769 | 130 | 5.25 |
| 40-44 | 1668 (11.87) | 1584 | 84 | 3.53 |
| 45-49 | 1338 (8.33) | 1268 | 70 | 4.59 |
| 50-54 | 1028 (6.42) | 968 | 60 | 4.3 |
| 55-59 | 713 (3.64) | 678 | 35 | 3.26 |
| 60-64 | 525 (2.31) | 500 | 25 | 2.97 |
| 65+ | 383 (1.69) | 358 | 25 | 4.62 |
| **Gender** |  |  |  |  |
| Female | 8633 (50.56) | 7976 | 657 | 6.06 |
| Male | 5741 (49.44) | 5487 | 254 | 3.47 |
| **Residence** |  |  |  |  |
| Rural | 7155 (47.89) | 6681 | 474 | 5.28 |
| Urban | 7219 (52.11) | 6782 | 437 | 4.32 |
| **Education level** |  |  |  |  |
| None | 2149 (10.07) | 2000 | 149 | 4.77 |
| Primary | 6388 (45.54) | 5947 | 441 | 5.59 |
| Secondary | 4404 (32.01) | 4134 | 270 | 4.51 |
| Tertiary | 1433 (12.38) | 1382 | 51 | 2.53 |
| **Occupation** |  |  |  |  |
| Government employee | 1474 (13.11) | 1426 | 48 | 2.36 |
| Non-Govt employee | 1444 (10.96) | 1353 | 91 | 5.12 |
| Self-employed | 4604 (30.09) | 4324 | 280 | 4.61 |
| Non-paid/Retired | 6852 (45.83) | 6360 | 492 | 5.51 |
| **Marital Status** |  |  |  |  |
| Never married | 3766 (29.96) | 3519 | 247 | 4.84 |
| Currently married | 9595 (65.48) | 9028 | 567 | 4.43 |
| Formerly married | 1013 (4.56) | 916 | 97 | 9.42 |
| **Country** |  |  |  |  |
| Algeria | 5546 (62.56) | 5375 | 171 | 3.16 |
| Eswatini | 2383 (0.76) | 2151 | 232 | 9.16 |
| Malawi | 3250 (19.13) | 3011 | 239 | 6.89 |
| Zambia | 3195 (17.55) | 2926 | 269 | 8.09 |
| **Diabetes Mellitus** |  |  |  |  |
| No | 13877 (96.49) | 13005 | 872 | 4.76 |
| Yes | 497 (3.51) | 458 | 39 | 5.37 |
| **Hypertension** |  |  |  |  |
| No | 12409 (88.45) | 11668 | 741 | 4.52 |
| Yes | 1965 (11.55) | 1795 | 170 | 6.80 |
| **High salt uptake** |  |  |  |  |
| No | 11109 (76.29) | 10457 | 652 | 4.52 |
| Yes | 3265 (23.71) | 3006 | 259 | 5.63 |
| **Low fruit uptake** |  |  |  |  |
| No | 4492 (44.92) | 4221 | 271 | 4.2 |
| Yes | 9882 (71.43) | 9242 | 640 | 5.01 |
| **Low vegetable uptake** |  |  |  |  |
| No | 9982 (71.73) | 9354 | 628 | 4.20 |
| Yes | 4392 (28.27) | 4109 | 283 | 5.01 |
| **Physical Activity** |  |  |  |  |
| No | 11503 (78.61) | 10739 | 764 | 5.06 |
| Yes | 2871 (21.39) | 2724 | 147 | 3.75 |
| **Smoking history** |  |  |  |  |
| Never | 11769 (75.64) | 11015 | 754 | 4.84 |
| Previous | 1051 (9.78) | 1006 | 45 | 2.64 |
| Current | 1554 (14.58) | 1442 | 112 | 5.93 |
| **Alcohol** |  |  |  |  |
| No | 11503 (97.39) | 13090 | 854 | 4.62 |
| Yes | 2871 (2.61) | 373 | 57 | 10.88 |
| **Survey year** |  |  |  |  |
| 2014 | 2383 (0.76) | 2151 | 232 | 9.16 |
| 2016 | 5546 (62.56) | 5375 | 171 | 3.16 |
| 2017 | 6445 (36.68) | 5937 | 508 | 7.46 |

**TABLE 2:** Determinants of factors associated with suicidal ideation of respondents that participated in the WHO STEPwise survey: 2014-2019.

| Characteristics (14,374) | Crude |  | Adjusted |  |
| --- | --- | --- | --- | --- |
|  | OR (95%CI) | P-value | AOR (95%CI) | P-value |
| **Gender** |  |  |  |  |
| Female | 1.00 |  | 1.00 |  |
| Male | 0.56 (0.46-0.68) | <0.0001 | 0.61 (0.52-0.72) | <0.0001 |
| **Age Group** |  |  |  |  |
| 15-19 | 1.00 |  | 1.00 |  |
| 20-24 | 1.14 (0.74-1.77) | 0.55 |  |  |
| 25-29 | 0.86 (0.56-1.32) | 0.49 |  |  |
| 30-34 | 1.05 (0.67-1.64) | 0.84 |  |  |
| 35-39 | 1.02 (0.66-1.56) | 0.94 |  |  |
| 40-44 | 0.67 (0.41-1.12) | 0.12 |  |  |
| 45-49 | 0.88 (0.50-1.56) | 0.67 |  |  |
| 50-54 | 0.83 (0.48-1.42) | 0.49 |  |  |
| 55-59 | 0.62 (0.35-1.09) | 0.10 |  |  |
| 60-64 | 0.56 (0.27-1.19) | 0.13 |  |  |
| 65+ | 1.00 |  | 1.00 |  |
| **Country** |  |  |  |  |
| Algeria | 1.00 |  | 1.00 |  |
| Eswatini | 3.09 (2.27-4.21) | <0.0001 | 3.24 (2.61-4.02) | <0.0001 |
| Malawi | 2.27 (1.62-3.20) | <0.0001 | 2.29 (1.83-2.85) | <0.0001 |
| Zambia | 2.70 (2.02-3.61) | <0.0001 | 2.66 (2.15-3.29) | <0.0001 |
| **Resistance** |  |  |  |  |
| Rural | 1.00 |  | 1.00 |  |
| Urban | 0.81 (0.62-1.06) | 0.12 |  |  |
| **Education level** |  |  |  |  |
| Primary | 1.181 (0.82-1.10) | 0.37 | 1.16 (0.95-1.42) | 0.15 |
| Secondary | 0.94 (0.64-1.38) | 0.76 | 1.03 (0.83-1.28) | 0.80 |
| Tertiary | 0.52 (0.31-0.87) | 0.01 | 0.72 (0.50-1.03) | 0.07 |
| **Occupation** |  |  |  |  |
| Non-Govt employee | 2.23 (1.30-3.85) | <0.0001 | 1.28 (0.87-1.86) | 0.21 |
| Self-employed | 2.00 (1.26-3.17) | <0.0001 | 0.97 (0.69-1.37) | 0.86 |
| Non-paid/Retired | 2.41 (1.56-3.70) | <0.0001 | 1.20 (0.85-1.69) | 0.30 |
| **Marital status** |  |  |  |  |
| Currently married | 0.91 (0.72-1.15) | 0.43 | 0.878 (0.74-1.04) | 0.13 |
| Formally married | 2.04 (1.44-2.91) | <0.0001 | 1.18 (0.90-1.53) | 0.23 |
| **Diabetes Mellitus** |  |  |  |  |
| No | 1.00 |  | 1.00 |  |
| Yes | 1.13 (0.68-1.89) | 0.63 |  |  |
| **Hypertension** |  |  |  |  |
| Yes | 1.54 (1.21-1.97) | <0.0001 | 1.50 (1.25-1.80) | <0.0001 |
| No | 1.00 |  | 1.00 |  |
| **High salt uptake** |  |  |  |  |
| Yes | 1.26 (1.03-1.53) | 0.02 | 1.38 (1.18-1.61) | <0.0001 |
| No | 1.00 |  | 1.00 |  |
| **Low fruit uptake** |  |  |  |  |
| Yes | 1.20 (0.96-1.51) | 0.11 |  |  |
| No | 1.00 |  | 1.00 |  |
| **Low vegetable uptake** |  |  |  |  |
| Yes | 1.10 (0.87-1.38) | 0.43 |  |  |
| No | 1.00 |  |  |  |
| **Alcohol** |  |  |  |  |
| Yes | 2.52 (1.67-3.82) | <0.0001 |  |  |
| No | 1.00 |  | 1.00 |  |
| **Physical Activity** |  |  |  |  |
| No | 1.00 |  | 1.00 |  |
| Yes | 1.00 |  | 1.00 |  |

**TABLE 3:** Characteristics and prevalence of individuals who took part in the WHO STEPwise survey on Suicidal Attempts from 2014-2019.

| Characteristics | Total | Suicidal Attempts |  |  |
| --- | --- | --- | --- | --- |
|  | N (%) | YES | NO |  |
| Social-demographic | 16,041 (100) | N | N | % |
| **Age Group** |  |  |  |  |
| 15-19 | 979 (7.31) | 918 | 61 | 5.16 |
| 20-24 | 1943 (16.36) | 1801 | 142 | 5.85 |
| 25-29 | 1891 (15.94) | 1758 | 133 | 4.47 |
| 30-34 | 2007 (13.51) | 1861 | 146 | 5.39 |
| 35-39 | 1899 (12.63) | 1769 | 130 | 5.25 |
| 40-44 | 1668 (11.87) | 1584 | 84 | 3.53 |
| 45-49 | 1338 (8.33) | 1268 | 70 | 4.59 |
| 50-54 | 1028 (6.42) | 968 | 60 | 4.3 |
| 55-59 | 713 (3.64) | 678 | 35 | 3.26 |
| 60-64 | 525 (2.31) | 500 | 25 | 2.97 |
| 65+ | 383 (1.69) | 358 | 25 | 4.62 |
| **Gender** |  |  |  |  |
| Female | 8633 (50.56) | 7976 | 657 | 6.06 |
| Male | 5741 (49.44) | 5487 | 254 | 3.47 |
| **Residence** |  |  |  |  |
| Rural | 7155 (47.89) | 6681 | 474 | 5.28 |
| Urban | 7219 (52.11) | 6782 | 437 | 4.32 |
| **Education level** |  |  |  |  |
| None | 2149 (10.07) | 2000 | 149 | 4.77 |
| Primary | 6388 (45.54) | 5947 | 441 | 5.59 |
| Secondary | 4404 (32.01) | 4134 | 270 | 4.51 |
| Tertiary | 1433 (12.38) | 1382 | 51 | 2.53 |
| **Occupation** |  |  |  |  |
| Government employee | 1474 (13.11) | 1426 | 48 | 2.36 |
| Non-Govt employee | 1444 (10.96) | 1353 | 91 | 5.12 |
| Self-employed | 4604 (30.09) | 4324 | 280 | 4.61 |
| Non-paid/Retired | 6852 (45.83) | 6360 | 492 | 5.51 |
| **Marital Status** |  |  |  |  |
| Never married | 3766 (29.96) | 3519 | 247 | 4.84 |
| Currently married | 9595 (65.48) | 9028 | 567 | 4.43 |
| Formerly married | 1013 (4.56) | 916 | 97 | 9.42 |
| **Country** |  |  |  |  |
| Algeria | 5546 (62.56) | 5525 | 21 | 0.37 |
| Eswatini | 2383 (0.76) | 2339 | 44 | 2.98 |
| Malawi | 3250 (19.13) | 3233 | 17 | 0.87 |
| Zambia | 3195 (17.55) | 3159 | 36 | 1.89 |
| **Diabetes Mellitus** |  |  |  |  |
| No | 13877 (96.49) | 13005 | 872 | 4.52 |
| Yes | 497 (3.51) | 458 | 39 | 6.8 |
| **Hypertension** |  |  |  |  |
| No | 12409 (88.45) | 11668 | 741 | 4.517 |
| Yes | 1965 (11.55) | 1795 | 170 | 6.80 |
| **High salt uptake** |  |  |  |  |
| No | 11109 (76.29) | 10457 | 652 | 4.52 |
| Yes | 3265 (23.71) | 3006 | 259 | 5.63 |
| **Low fruit uptake** |  |  |  |  |
| No | 4492 (28.57) | 4221 | 271 | 4.2 |
| Yes | 9882 (71.43) | 9242 | 640 | 5.01 |
| **Low vegetable uptake** |  |  |  |  |
| No | 9982 (71.73) | 9354 | 628 | 4.66 |
| Yes | 4392 (28.27) | 4109 | 283 | 5.09 |
| **Physical Activity** |  |  |  |  |
| No | 11503 (78.61) | 10739 | 764 | 5.06 |
| Yes | 2871 (21.39) | 2724 | 147 | 3.75 |
| **Smoking history** |  |  |  |  |
| Never | 11769 (75.64) | 11015 | 754 | 4.84 |
| Previous | 1051 (9.78) | 1006 | 45 | 2.64 |
| Current | 1554 (14.58) | 1442 | 112 | 5.93 |
| **Alcohol** |  |  |  |  |
| No | 11503 (97.39) | 13090 | 854 | 4.62 |
| Yes | 2871 (2.61) | 373 | 57 | 10.88 |
| **Survey ear** |  |  |  |  |
| 2014 | 2383 (0.76) | 2151 | 232 | 9.16 |
| 2016 | 5546 (62.56) | 5375 | 171 | 3.16 |
| 2017 | 6445 (36.68) | 5937 | 508 | 7.46 |

**TABLE 4:** Determinants of factors linked with suicidal Attempts among participants in the WHO STEPwise survey from 2014-2019.

| Characteristics | Crude |  | Adjusted |  |
| --- | --- | --- | --- | --- |
|  | OR (95%CI) | P-value | AOR (95%CI) | P-value |
| **Gender** |  |  |  |  |
| Female | 1.00 |  | 1.00 |  |
| Male | 0.56 (0.46-0.68) | <0.0001 | 0.61 (0.52-0.72) | <0.0001 |
| **Age** |  |  |  |  |
| 15-19 | 1.00 |  | 1.00 |  |
| 20-24 | 1.14 (0.74-1.77) | 0.55 |  |  |
| 25-29 | 0.86 (0.56-1.32) | 0.49 |  |  |
| 30-34 | 1.05 (0.67-1.64) | 0.84 |  |  |
| 35-39 | 1.017 (0.66-1.56) | 0.94 |  |  |
| 40-44 | 0.673 (0.41-1.12) | 0.12 |  |  |
| 45-49 | 0.884 (0.50-1.56) | 0.67 |  |  |
| 50-54 | 0.825 (0.48-1.42) | 0.49 |  |  |
| 55-59 | 0.618 (0.35-1.09) | 0.10 |  |  |
| 60-64 | 0.56 (0.27 1.19) | 0.13 |  |  |
| 65+ | 1.00 |  |  |  |
| **Country** |  |  |  |  |
| Algeria | 1.00 |  | 1.00 |  |
| Eswatini | 3.09 (2.27-4.21) | <0.0001 | 3.24 (2.61-4.02) | <0.0001 |
| Malawi | 2.273 (1.62-3.20) | <0.0001 | 2.29 (1.83-2.85) | <0.0001 |
| Zambia | 2.70 (2.02-3.61) | <0.0001 | 2.66 (2.15-3.29) | <0.0001 |
| **Resistance** |  |  |  |  |
| Rural | 1.00 |  | 1.00 |  |
| Urban | 0.81 (0.62-1.06) | 0.12 |  |  |
| **Education level** |  |  |  |  |
| Primary | 1.181 (0.82-1.10) | 0.37 | 1.16 (0.95-1.42) | 0.15 |
| Secondary | 0.94 (0.64-1.38) | 0.76 | 1.03 (0.83-1.28) | 0.80 |
| Tertiary | 0.52 (0.31-0.87) | 0.01 | 0.72 (0.50-1.03) | 0.07 |
| **Occupation** |  |  |  |  |
| Non-Govt employee | 2.23 (1.30-3.85) | <0.0001 | 1.28 (0.87-1.86) | 0.21 |
| Self-employed | 2.00 (1.26-3.17) | <0.0001 | 0.97 (0.69-1.37) | 0.86 |
| Non-paid/Retired | 2.41 (1.56-3.70) | <0.0001 | 1.20 (0.85-1.69) | 0.30 |
| **Marital status** |  |  |  |  |
| Currently married | 0.91 (0.72-1.15) | 0.43 | 0.878 (0.74-1.04) | 0.13 |
| Formally married | 2.04 (1.44-2.91) | <0.0001 | 1.18 (0.90-1.53) | 0.23 |
| **Diabetes (dbm)** |  |  |  |  |
| No | 1.00 |  | 1.00 |  |
| Yes | 1.13 (0.68-1.89) | 0.63 |  |  |
| **Hypertension (bp)** |  |  |  |  |
| Yes | 1.54 (1.21-1.97) | <0.0001 | 1.50 (1.25-1.80) | <0.0001 |
| No | 1.00 |  | 1.00 |  |
| **High salt uptake** |  |  |  |  |
| Yes | 1.26 (1.03-1.53) | 0.02 | 1.38 (1.18-1.61) | <0.0001 |
| No | 1.00 |  | 1.00 |  |
| **Low fruit uptake** |  |  |  |  |
| Yes | 1.20 (0.96-1.51) | 0.11 |  |  |
| No | 1.00 |  | 1.00 |  |
| **Low vegetable uptake** |  |  |  |  |
| Yes | 1.10 (0.87-1.38) | 0.43 |  |  |
| No | 1.00 |  |  |  |
| **Alcohol** |  |  |  |  |
| Yes | 2.52 (1.67-3.82) | <0.0001 |  |  |
| No | 1.00 |  | 1.00 |  |
| **Physical Activity** |  |  |  |  |
| No | 1.00 |  | 1.00 |  |
| Yes | 1.00 |  | 1.00 |  |

## DISCUSSION

This study assessed suicidal ideation and suicidal attempts among over 14000 participants across Algeria, Malawi, Zambia and Eswatini. Overall, 6.34% of individuals reported suicidal ideation and 5.69% reported suicide attempts. Females, younger age groups and individuals with low educational attainment showed higher prevalence. Behavioral and biomedical factors especially hypertension, alcohol use and high salt intake were strongly linked to increased suicide risk, Geographic disparities were striking with Eswatini showing the highest adjusted odds for suicidal thoughts and attempts, followed by Zambia and Malawi. Contextual factors such as country of residence proved to be stronger predictors than demographic variables like age, marital status or occupation once adjustments were made. Importantly, alcohol use stood out as a critical behavioral determinant, with nearly double the risk for those who consumed alcohol. Other lifestyle factors, such as low fruit and vegetable intake as well as physical inactivity showed weaker associations. The findings underscore the complex interplay of social, behavioral and health-related factors shaping suicidal ideation and risk across different settings.

Globally, women report higher rates of suicidal ideation, while men are more likely to die by suicide due to the use of more lethal means (32,33). This study aligns with global trends in ideation but diverges on attempts, as women were also more likely to attempt suicide. In the African context, this may reflect gender-specific vulnerabilities such as economic dependence, gender-based violence, and limited access to mental health care for women (34). These findings highlight the urgent need for gender-sensitive mental health interventions, including the integration of mental health services into reproductive and maternal health programs, expanded access to psycho-social support, and the scaling up of safe spaces and helplines for women (35,36).

The significantly higher odds of suicidal ideation and attempts in Eswatini, Malawi, and Zambia compared to Algeria point to the critical role of socio-political and economic contexts. Factors such as high poverty rates, HIV prevalence, and weak mental health systems particularly in Eswatini and Malawi may contribute to elevated psychological distress (37). These findings underscore the need for country-specific mental health strategies. Policymakers should prioritize national suicide prevention plans that incorporate community-based interventions, mental health workforce training, and integration of services into primary care, especially in resource-limited settings (38,39).

The association between suicidal behaviors and hypertension or high salt intake highlights the interplay between physical and mental health (40). Hypertension may act as a stressor due to chronic disease burden or medication side effects, while high salt intake has been linked with mood disregulations in emerging biomedical research (41). As non-communicable diseases rise across Africa, these findings call for holistic health policies. Public health efforts should adopt integrated care models that include mental health screening for NCD patients, dietary counseling, and training clinicians to identify psychological distress in individuals with hypertension (42),(43).

Tertiary education was protective against suicidal ideation and attempts, consistent with global evidence linking education to greater coping capacity and resource access (44). Conversely, individuals with low or no formal education often in rural African communities face structural disadvantages, including unemployment and limited mental health literacy (45). These findings call for equity-focused mental health policies that promote education and employment. Governments should invest in community awareness campaigns, strengthen school-based mental health programs, and expand social safety nets for vulnerable, low-education populations(46),(47).

Although not included in adjusted models, alcohol use showed a strong crude association with suicide attempts. Globally, alcohol is a well-established risk factor for suicidal behavior, and in SSA, weak regulation exacerbates its role in impulsivity and poor mental health (48,49). The policy implications are clear: governments should implement stricter alcohol control measures such as taxation, restrictions on unrecorded alcohol, and community-based harm-reduction strategies to promote healthier coping mechanisms (50).

A key strength of this study is its large, multi-country sample from diverse African settings, enabling cross-country comparisons and regionally relevant insights (51). The use of adjusted odds ratios strengthens the validity of associations by accounting for potential confounders, while the inclusion of both demographic and biomedical variables offers a comprehensive perspective on suicide risk factors (52). Standardized survey methods further enhance internal validity and comparability across countries (53).

However, several limitations should be noted. The cross-sectional design precludes causal inference, as temporal relationships cannot be established. Self-reported data may be affected by recall and social desirability bias, particularly for sensitive topics like suicide and alcohol use, potentially leading to under reporting. Additionally, the absence of adjusted models for some variables such as alcohol use limits interpretation of their independent effects. Despite these limitations, the study provides valuable evidence for policy planning and underscores the need for complementary longitudinal and qualitative research to explore causal pathways and inform suicide prevention strategies in SSA.

In conclusion, this study reveals a substantial burden of suicidal ideation and attempts in several African countries, particularly among females, individuals with hypertension, those consuming high-salt diets, and residents of Eswatini, Malawi, and Zambia. The findings highlight the urgent need to integrate mental health services into primary healthcare systems, with a focus on high-risk groups. In resource-limited settings where mental health infrastructure is often weak, governments should prioritize community-based mental health promotion, expand access to psycho-social support, and strengthen early detection through routine screening. Integrating mental health education into schools, workplaces, and community programs can reduce stigma and improve help-seeking behaviors. Integrating mental health education into schools, workplaces, and community programs can reduce stigma and improve help-seeking behaviors. Policymakers must also invest in training the mental health workforce and ensuring that services are culturally appropriate, affordable, and accessible These actions are essential not only to reduce suicide rates but also to enhance overall public health and well-being in Africa and other low-resource settings .m

## Data Availability

ALL The datasets were accessed via the WHO NCD Micro-data Repository at https://extranet.who.int/ncdsmicrodata/index.php/home

## AUTHOR CONTRIBUTIONS

Conceptualization I.N., (Ilunga Ngalande), A.C (Adoration Chigere), W.N (Wingston Ng’ambi) and C.Z (Cosmas Zyambo); methodology, I.N. (Ilunga Ngalande) and (Wingston Ng’ambi); formal analysis, I.N., (Ilunga Ngalande); investigation, I.N., (Ilunga Ngalande); writing original draft preparation, I.N., (Ilunga Ngalande) and A.C (Adoration Chigere); writing review and editing, (Ilunga Ngalande), A.C (Adoration Chigere) and W.N (Wingston Ng’ambi) ; supervision, CZ and W.N (Wingston Ng’ambi); revisions, C.Z, A.C, and I.N., (Ilunga Ngalande); All authors have read and agreed to the published version of the manuscript

## Acknowledgments

The authors would like to acknowledge the participants for taking their time and valuable information, without them which this study couldn’t be successful.

